# The Clinical Genomic Variation Landscape

**DOI:** 10.1101/2025.11.04.25339115

**Authors:** Wesley A. Goar, Daniel Puthawala, Kori Kuzma, Anastasia Bratulin, Austin A. Antoniou, Jeremy A. Arbesfeld, Lawrence Babb, Kyle Ferriter, Terry O’Neill, James S. Stevenson, Kathryn Perry, Matthew Cannon, Jiachen Liu, Xuelu Liu, Brian Walsh, Savanna Funk, William C. Ray, Bimal P. Chaudhari, Heidi L. Rehm, Alex H. Wagner

**Affiliations:** The Steve and Cindy Rasmussen Institute for Genomic Medicine, Nationwide Children’s Hospital, Columbus, OH; The Ohio State University Biomedical Sciences Graduate Program, Columbus, OH; Medical and Population Genetics, Broad Institute of MIT and Harvard, Cambridge, MA; Dana Farber Cancer Institute, Boston, MA; Oregon Health and Science University, Portland, OR; The Abigail Wexner Research Institute Graphics Resource, Nationwide Children’s Hospital, Columbus, OH; The Ohio State University Biophysics Program, Columbus, OH; Center for Genomic Medicine, Massachusetts General Hospital, Boston, MA; Divisions of Neonatology, Genetic and Genomic Medicine, Department of Pediatrics, Nationwide Children’s Hospital, Columbus, OH; Departments of Pediatrics and Biomedical Informatics, The Ohio State University College of Medicine, Columbus, OH; Clinical and Translational Sciences Institute, Nationwide Children’s Hospital and The Ohio State University, Columbus, OH

**Author notes:** These authors contributed equally to this work (Wesley A. Goar, Daniel Puthawala).

## Abstract

Interpreting genomic variation requires analysts to collate and process information from disparate genomic evidence resources to discern the contributions to diseases and drug responses. Differences in variant representation across these evidence repositories includes nomenclature (e.g., HGVS, SPDI), reference sequence context (e.g., GRCh37 or GRCh38 genome assemblies), sequence annotation sources (e.g., RefSeq or Ensembl), and aggregate variant concepts (e.g., canonical alleles) collectively make it difficult to reveal whether (and how) genomic variants are associated with clinical outcomes. We evaluated these challenges across established genomic knowledge resources, including content from the CIViC, Molecular Oncology Almanac, and ClinVar knowledgebases, as compared against real-world small variant and CNV data. We used these findings to develop a suite of variant normalization methods to address these gaps. We present our findings as well as an analysis of remaining gaps in the representation of variation data and recommendations for the continued development of genomic knowledge standards to address these gaps.

## Introduction

Genomic medicine has rapidly emerged as a transformative force in health care, driven by remarkable advancements in sequencing technologies. Integral to the practice of genomic medicine is variant interpretation, which requires collection of multiple lines of evidence from genomic evidence repositories and knowledgebases to support or refute the clinical significance of the assayed genomic variants. However, disparate representations of variants across resources create inconsistencies that hinder interoperability between resources^1,2^. Therefore, clinical variant scientists, clinicians, and curators must undertake the laborious task of manually collecting and interpreting evidence from multiple sources for application within clinical workflows^3–6^. This effort becomes a rate-limiting step to the realization of scalable genomic medicine, and is known as the *interpretation bottleneck*^7^. The interpretation bottleneck presents unique challenges due to the inherent complexity of genomic knowledge and the many ways in which it can be represented. For variant interpretation (and reinterpretation) to become automatic, scalable, and reproducible in the practice of genomic medicine, novel computational standards and supporting data harmonization tools that address this bottleneck are required.

Several factors contribute to the complexity of managing variant representation across genomic resources^8^ (**Figure 1**). Changes to reference sequences between human genome builds create different coordinate systems, leading to shifts in the alignment of variants called against different builds over time (i.e. GRCh37 vs. GRCh38), and thereby complicating the reconciliation of evidence (**Figure 1A**). Additionally, transcript selection may affect variant interpretation, as genomic variants projected onto a given gene transcript isoform sequence may result in different coordinate positions or be altogether absent when projected onto another transcript sequence from the same gene, resulting in significantly different representations–and possibly interpretations–of variant effect (**Figure 1B**).

**Figure 1.**
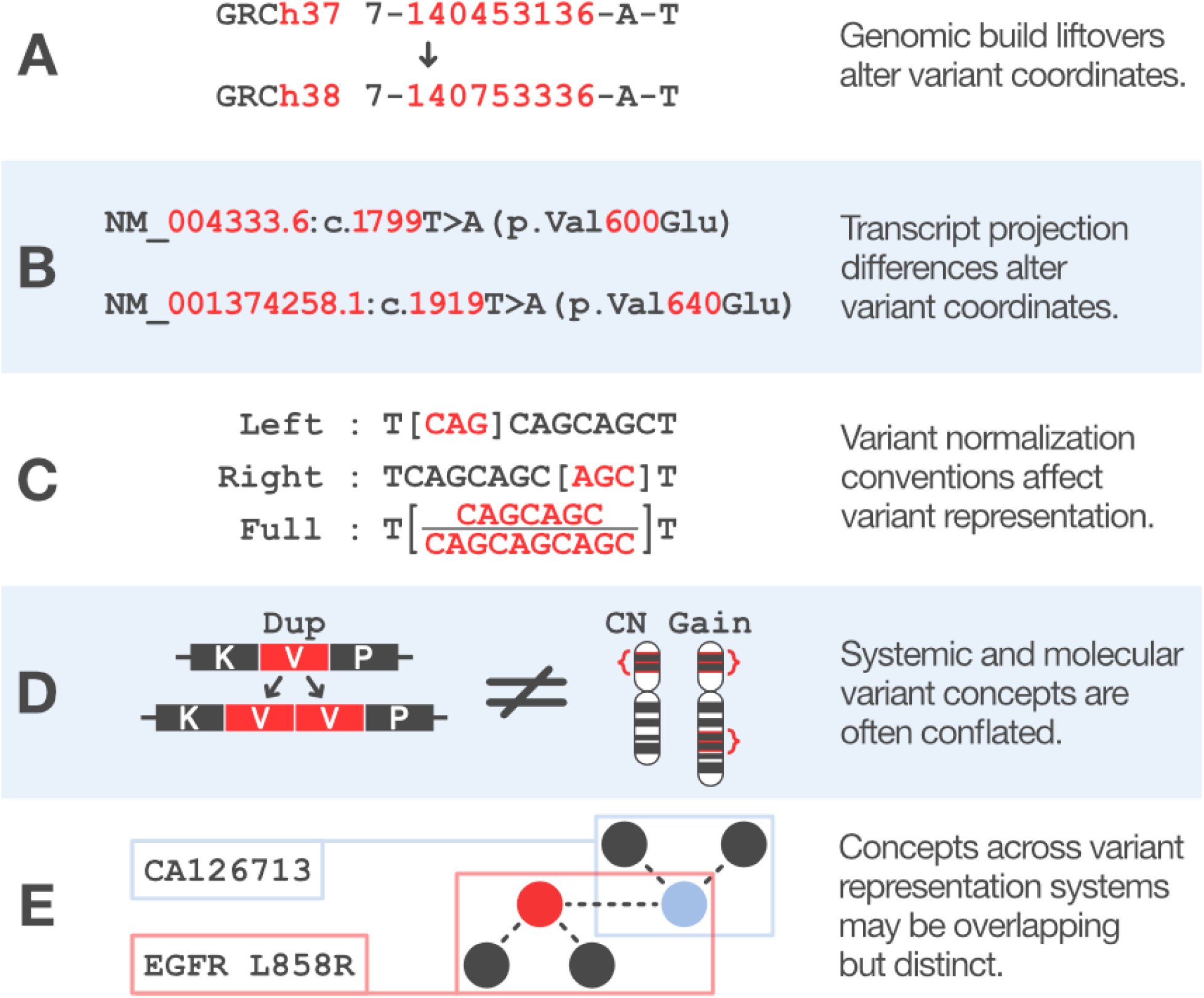
Challenges to harmonizing variant representation across genomic resources. There are a number of challenges to variant normalization across genomic resources. These include variant coordinate changes resulting from differences in **(A)** reference genome build or **(B)** choice in transcript being projected to. **(C)** Within a given sequence, differing normalization conventions can non-trivially alter how a given underlying sequence can be represented. **(D)** Different nomenclatures and variant representation systems can also cause conflicts when distinct variant concepts are conflated, or **(E)** partially overlap.

Sequence variants are also often represented using standard nomenclatures (e.g. HGVS^9^, SPDI^10^), or resource-specific identifiers derived from these or related variant data standards (e.g. VCF^11^). When describing a variant using a nomenclature, conventions are needed for normalizing ambiguous insertions or deletions (indels) occurring in repeating regions, as the exact location of these indels cannot be precisely determined and different standards will use different conventions for resolving these (**Figure 1C**). Often, resources are unable to prevent community (ab)use of variant nomenclatures to describe variant forms the nomenclature does not support, leading to downstream misinterpretation of the variant. A common example of this is expressing systemic copy number events (the relative abundance of a genomic subsequence in a sample^12^) using the HGVS nomenclature tandem duplication syntax (a duplication event occurring in tandem on a contiguous sequence) (**Figure 1D**).

Representations of specific variants across nomenclatures remains an unsolved problem; conceptually overlapping yet non-identical variants may be represented using incompatible identifiers (e.g., EGFR L858R vs CA126713), introducing further challenges to accurate variant curation and clinical interpretation (**Figure 1E**). In addition to standard variant nomenclatures, knowledgebases may also utilize registered identifiers such as dbSNP^13^, rsIDs, or ClinGen Allele Registry IDs^14^; computed identifiers^12^; and/or structured variant representations using separated data fields (e.g. start/end/ref/alt)^8^. Resource-specific nomenclatures and naming conventions also commonly appear across knowledgebases, in part due to a lack of widely-accepted standards for describing *categorical variants*^1,15^–variant concepts described by properties of variants (e.g. “MET exon 14 skipping mutations”) to which other assayed variants may match. Categorical variants of this type are commonly found in somatic cancer knowledgebases and clinical trial criteria, though *canonical alleles* (a class of categorical variation) are also frequently used in clinical resources^10,14^.

To better understand the effect of these challenges on automated search and retrieval of genomic evidence, we assess the feasibility of normalizing genomic variants from real-world clinical cohorts using genomic evidence from the Clinical Interpretation of Variants in Cancer (CIViC), Molecular Oncology Almanac^16^ (MOAlmanac), and ClinVar knowledgebases. We highlight specific challenges we encountered (**Figure 1**) and describe the Variation Normalizer (normalize.cancervariants.org/variation), an open-source toolkit we developed to address those challenges, on small variant data and copy number variant (CNV) data from multiple real-world patient cohorts and knowledgebases. We categorize common classes of variants that cannot be normalized using current data standards, and the expected clinical impact of resolving these representation gaps. We use these findings to create a prioritized list of variant categories that, once normalized, have the greatest potential to impact clinical actionability.

Around the world, genomic data analyses rely upon re-engineering solutions to handle the complex landscape of variant representation across clinical assays and knowledge resources. This study identifies key challenges commonly addressed in these data harmonization efforts, and provides open-source tools to address those challenges. Our approach provides tools for the systematic harmonization of genomic variant types common to many research and clinical genomics workflows today, and a community roadmap for challenges yet to be addressed.

## Results

### Normalization of variants and knowledge matching to real-world data

Annotating patient variants with knowledge significant to clinical outcomes and therapeutic interventions are foundational practices within genomic medicine. We evaluated methods for matching clinical evidence from knowledgebases to real world data, comparing somatic variant data from the American Association for Cancer Research’s (AACR) Project Genomics Evidence Neoplasia Information Exchange (GENIE)^17^ and germline CNV data from Nationwide Children’s Hospital (NCH). Using the Variation Normalizer, we created normalized, computable representations of the genomic and protein representations of variants (**Figure 2A**) to computable objects from these datasets, as well as variant data from the CIViC, MOAlmanac, and ClinVar knowledgebases. We then searched the knowledgebase content with observed variants from the cohort data (**Figure 2B, Supplemental Table 1A)** as an evaluation of our ability to match real world data to clinical evidence.

**Figure 2.**
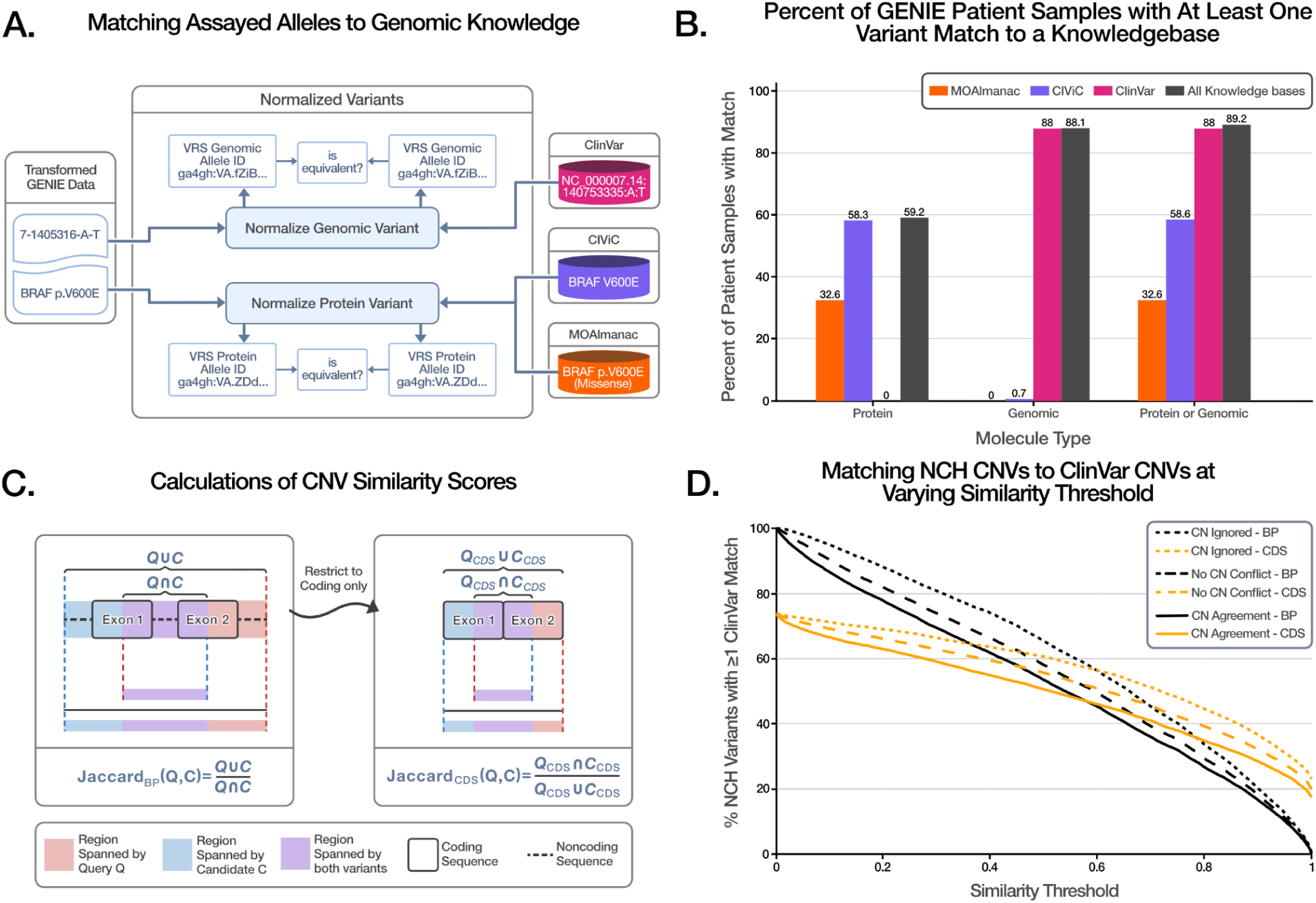
Generalizable strategies for matching small variants and CNVs to clinical knowledgebases. **(A)** We applied a variant harmonization and match strategy for genomic and protein alleles represented in different syntaxes from the MOAlmanac, CIViC, and ClinVar knowledgebases, which we compared to the and AACR Project GENIE dataset using computed VRS IDs. **(B)** Standardized representation via VRS allows patient variants from GENIE to be matched to variants from the MOAlmanac, CIViC, and ClinVar knowledgebases, which facilitates downstream interpretation. **(C)** Matching Patient CNVs to ClinVar CNVs is rarely an exact match, and so Jaccard scores were calculated to assess degrees of matching. The Jaccard score is equal to the set intersection divided by the set union. **(D)** We computed scores for both all base pairs (BP, in black) in the CNV and for just the coding sequence (CDS, in orange), and plotted the percentage of CNVs that match as the Jaccard score threshold increases from almost no overlap to 1, a perfect match, for both the BP and CDS scores. Additionally, we represented several degrees of strictness of copy number match: *CN Ignored* (dotted line) did not consider either CNV’s copy count; *No CN Conflict* (dashed line) meant that the copy numbers for both CNVs did not disagree (e.g. one’s copy number is a range which included the other’s copy number); and *CN agreement* (solid line) required patient and ClinVar CNVs copy numbers to be equal.

There were a total of 204,292 unique patient samples in the GENIE data set, with an average of 13.41 variants per sample. We successfully normalized an average of 12.26 variants per patient sample; and successfully normalized at least one variant for 98% (199,835/204,292) of the samples. Using the Variation Normalizer, we found that 91% (182,128/199,835) samples in GENIE with at least one normalized variant could be matched to knowledge in at least one of the three clinical interpretation knowledgebases (**Figure 2B**). Of all samples, 59% (119,814) had a normalized variant that matched to knowledge in CIViC, 33% (66,641) matched to knowledge in MOAlmanac, and 88% (179,862) matched to knowledge in the ClinVar knowledgebase (**Figure 2B**). For samples with at least one variant, we analyzed the average number of normalized variants per patient sample, stratified by genomic and protein representations. We found that an average of 10.79 normalized protein variants and 12.08 normalized genomic variants per sample, and found that on average 1.46 of these normalized variants matched to clinical evidence within CIViC, 1.14 matched to MOAlmanac, and 5.59 matched to ClinVar (**Supplemental Table 1B**).

We extracted 19,106 (14,710 unique) CNVs from the text of microarray reports of tests performed at a Nationwide Children’s Hospital (see **Methods**). We excluded 27 malformed ISCN strings, 5,683 regions of homozygosity, and 11 variants in GRCh36, after which 8,989 variants remained. The Variation Normalizer was able to normalize 97% (8,718/8,989) of these variants. Of the 271 which could not be normalized, 63 failed due to invalid sequence positions, and 208 were unable to be lifted over to GRCh38 (**Supplemental Figure 1B**).

We determined which NCH CNVs had a matching ClinVar CNV using a range of criteria, based on simple base pair (BP) overlap or coding sequence (CDS) overlap (**Figure 2C**). Exact matches under BP overlap—BP Jaccard score equal to one—were exceedingly rare, realized only for 0.2% (20/8,718) of NCH CNVs (**Figure 2D**, rightmost point on dotted black curve). Even relaxing the Jaccard similarity threshold score to 0.9 only admitted matches for 20.1% (1,750/8,718) of NCH variants, without taking into account agreement/disagreement of copy numbers between NCH-ClinVar CNV pairs (**Figure 2D**, x=0.9 on dotted black curve). By comparison, matching CNVs by the CDS regions they overlap was more forgiving, realizing matches for 17.3% (1,511/8,718) of NCH CNVs under the strictest CDS matching: CDS Jaccard score equal to one, with the additional restriction that the CNVs’ copy number information must be compatible (**Figure 2D**, rightmost point on solid orange curve).

### Tools to enable open source variant normalization in genomic medicine

We developed our variant normalization methods through the Variation Normalizer^18^ (**Figure 3**), an open-source Python package and REST API accessible online at normalize.cancervariants.org/variation. This open-source software leverages FastAPI^19^ and the OpenAPI specification, lowering barriers to interoperability with modern clinical variant interpretation workflows. The Variation Normalizer provides methods for the translation of molecular and systemic variation represented in a free-text format to computable data objects following the Variation Representation Specification^12,20^ (VRS) format of the Global Alliance for Genomics and Health (GA4GH)^21^. The software is open-source, and the codebase is broadly licensed (MIT license) for reuse in any clinical and research genomics workflows.

**Figure 3.**
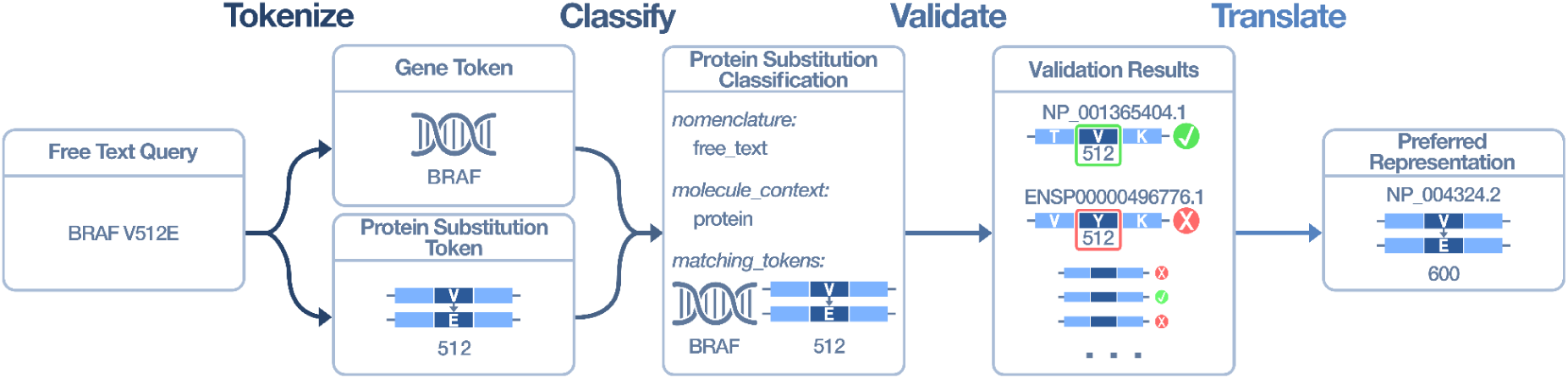
The Variation Normalizer converts free-text representations of variants into standardized computable objects. In this example, we see the free text query “BRAF V512E” converted to a computable object representing NP_004324.2:p.Val600Glu, through four discrete steps. **(Tokenize)** Pattern matching is used to split text strings into conceptual entities. **(Classify)** Combinations of conceptual entities are used to classify the variant type, such as a Protein Substitution variant. **(Validate)** The variant content is validated against reference sequences to identify variant contexts that are concordant with the queried reference location and residue. **(Translate)** A computable object is created following preferred representation conventions (e.g. on GRCh38 or a protein sequence derived from a MANE Select transcript.

The Variation Normalizer takes a query as input and performs four steps to return a VRS object (**Figure 3**). During the first step, *tokenization*, the query is split into discrete conceptual entities (tokens) such as gene or allele. *Classification* performs exact pattern matching on the ordered tokens to determine the conceptual type of the variant, authenticates the nomenclature syntax, and collects additional properties relevant to the query such as nomenclature and molecular context. The *validation* step retrieves overlapping sequence accessions (chromosome, transcript, and/or protein sequences) for the classification and verifies that the expected reference sequence matches an actual sequence for a given accession at a given position (e.g., *Is there a Valine amino acid at position 512 on this reference sequence?*). Finally, during *translation*, results are lifted over to the GRCh38 build (if on GRCh37), a preferred transcript is selected (e.g., MANE v1.4), the Allele is normalized following the VRS conventions^12^, and the conceptual information is exported to the VRS data model. Currently supported query formats include: HGVS (e.g., NM_004333.6:c.1799T>A), VCF-strings as seen in resources like gnomAD^22^ (e.g., 7-140753336-A-T), and common free-text expressions as typically found in resources like CIViC^4^ (e.g., BRAF V600E).

The Variation Normalizer can translate both assayed variant data and variants found in clinical genomic knowledgebases. Users can represent CNVs using input HGVS copy number expressions with a precise copy number count, a copy number change (e.g., gain or loss), or parsed components (e.g., chromosome, start position, and end position). For the NCH and ClinVar CNV analysis, exact positions were used if available, otherwise, inner positions (see methods) were prioritized. A combination of precise copy number count (e.g., x3) and copy number change (e.g., gain or loss) expressions was also utilized. The Variation Normalizer exposes Cool-Seq-Tool’s^23^ *feature_overlap* endpoint in the REST API to find overlapping MANE^24^ features (gene and CDS) given GRCh38 genomic data (chromosome, start position, end position). Finally, the start and stop for CDS regions and overlap regions are defined using VRS Sequence Locations.

### Variation Normalizer Performance & Knowledgebase Analysis

#### CIViC Normalization Performance

At the time of our analysis, the CIViC database contained 3,845 variants whose associated entities matched an “accepted” or “submitted” status. The Variation Normalizer successfully normalized 52% (2,015/3,845) of variants and was unable to normalize 2% (83/3,845) of variants. The remaining 45% (1,747/3,845) of variants were outside the scope of this initial version of the Variation Normalizer and so were not supported (**Supplemental Table 2A**). 83% (69/83) of variants that were unable to be normalized were of the status “submitted” - variants that had not yet been reviewed by the CIViC curators^25^. After our manual review, all of these variant entries either followed non-standard nomenclature practices, or referred to a combination of positions and amino acids that don’t exist on a supported transcript such as “BRAF V601E”. These records represent the complexity of variant alias curation that require additional human intervention to contextualize or re-curate under standard nomenclature conventions.

All variants analyzed were binned under a common category schema based on the category creation analysis (see **methods** for category definitions). The largest group of variants that we were unable to normalize were “transcript variants”, which represented 21% (362/1,747) of all failed normalization attempts. These were primarily variants whose names contain a “c.” reference sequence prefix with no genomic HGVS expression, and no representative coordinate to form a VCF-like expression, such as “*VHL* A56_P59del (c.166_178del)”. In these cases, no attempt was made to normalize them, as a “c.” HGVS-like expression in the name indicates that the variant is a transcript-contextualized genomic variant, and there is no community policy for preferred representation of indels spanning exon boundaries. The policy for handling ranged indels on transcripts is under development as a planned feature. The complete breakdown of unsupported variants for CIViC within their respective categories can be found in **Supplemental Table 2B**.

#### Molecular Oncology Almanac Normalization Performance

At the time of our analysis, MOAlmanac contained 452 variants. The majority of these (57%; 256/452) were of types not supported by Variation Normalizer. All of the remaining 196 variants successfully normalized (**Supplemental Table 3A**). The largest category of variants not supported by Variation Normalizer were classified as “sequence variants”, but only included the gene name without any specific variant level information such as “*TP53* somatic variant” or “*AKT2* somatic variant” (28%; 127 / 452). The next largest category of unsupported variants were “region-defined” variants which only specified a specific region, such as “*TP53* exon 2 somatic variant” (9%; 40 / 452). We classified these and the remaining unsupported variants from MOAlmanac across twelve variant categories, as summarized in **Supplemental Table 3B**.

#### ClinVar Normalization Performance

The July 29th, 2025 release of ClinVar contains 3,757,467 unique variants. The Variation Normalizer successfully normalized 99.67% (3,745,114/3,757,464) and failed to normalize 0.06% (2,118/3,757,464) of these variants. The remaining 0.27% (10,235/3,757,464) of variants are outside the scope of this initial version of the Variation Normalizer and so were excluded from the analysis. The vast majority of ClinVar variants were single nucleotide variants, of which 99.99% were (3,401,625/3,401,990) normalized. The next-largest group of variants were deletions, of which 99.56% (160,232/160,943) were normalized. Overall, the majority of variants that failed to normalize were deletions/CNVs. Some examples of failed CNVs were “t(1;3)(q21.1;p11.1)dn” or “NM_007294.3(*BRCA1*):c.1387_1390delAAAAins5”, which failed due to unsupported syntax. Another cause of normalization failure was an unsuccessful liftover to GRCh38. Most variants of unsupported types had no HGVS expression and location, had an invalid HGVS expression, or were only using NCBI build 36 genomic coordinates for which liftover wasn’t supported. The rest were genotypes or haplotypes which could have been modeled in VRS^12,20^ but were not supported by this initial version of the Variation Normalizer.

#### Combined Normalization Results

We assessed the performance of the Variation Normalizer in normalizing variant concepts across the three knowledgebases assessed in this study. Overall, the Variation Normalizer successfully normalized 99.39% of the 2,214,573 variants in CIViC, MOAlmanac, and ClinVar (Table 1). However, this performance is dominated by the application of the service to the (relatively much larger) ClinVar knowledgebase. We also looked more closely at the clinical impact of variants not supported in the somatic cancer knowledgebases, which rely heavily on the use of categorical variants^1,15^.

**Table 1.** Counts, Normalizer Performance, and Data Types of Variants by Category. Variants are grouped by high-level variant type categories and summarized across the aggregated variant content from the CIViC, MOAlmanac, and ClinVar knowledgebases. The percentage of those variants which normalized was also calculated. The rightmost six columns reflect variant properties that can be found in the respective knowledgebases. Boxes marked with a solid circle indicate properties that are required for variant entries of each respective variant category. Hollow circles mark properties that are not required, but are frequently found in variants for a given category.

Counts, Normalizer Performance, and Data Types of Variants by Category
| Variant Category | Count | % Normalized | Δ Sequence | Δ Location | Δ Frame | Δ Quantity | Δ Function | Region Specificity |
| --- | --- | --- | --- | --- | --- | --- | --- | --- |
| Sequence | 3,710,957 | 99.84% | ● | ○ | ○ | ○ | ○ | ○ |
| Genotype/Haplotype | 915 | 0.00% | ● | ○ | ○ | ○ | ○ | ○ |
| Fusion | 318 | 0.00% |  | ● |  |  |  | ○ |
| Rearrangement | 441 | 0.00% |  | ● |  |  |  | ○ |
| Epigenetic Modification | 14 | 0.00% |  |  |  | ● | ○ | ○ |
| Copy Number | 47,588 | 88.14% |  |  |  | ● | ○ | ○ |
| Expression | 305 | 0.00% |  |  |  | ○ | ● | ○ |
| Gene Function | 125 | 4.80% |  |  |  |  | ● | ○ |
| Region-Defined | 326 | 8.59% |  |  |  |  |  | ● |
| Genome Feature | 10 | 0.00% |  |  |  | ○ | ○ |  |
| Other | 742 | 51.21% | ○ | ○ | ○ | ○ | ○ | ○ |

● Core Information Fields
○ Optional Information Fields

Variants in CIViC and MOAlmanac contained similar evidence statement rating systems and many of these were closely related to the AMP/ASCO/CAP clinical actionability tiering system. We evaluated the putative clinical impact of evidence from unsupported variants in these knowledgebases, using evidence level scores from CIViC and MOAlmanac following a uniform scoring scale. Every evidence statement within each knowledgebase was assigned an impact score ranging from 0.5-10 (see **methods**). A score of 10 was associated with FDA-approved therapy data (MOAlmanac) or validated association data (CIViC) which are often already in routine clinical practice. For a variant to be scored, it needed to have at least one evidence item associated with it within CIViC (either “submitted” or “accepted”) or MOAlmanac. For a variant with multiple evidence items, the total impact score of that variant was equal to the sum of the impact scores across all evidence items. The overall category impact score was the sum of the individual variant impact scores for that given category. We used this combined category score to identify the most clinically meaningful categories of variants to support in future versions of the Variation Normalizer (**Figure 4**).

**Figure 4:**
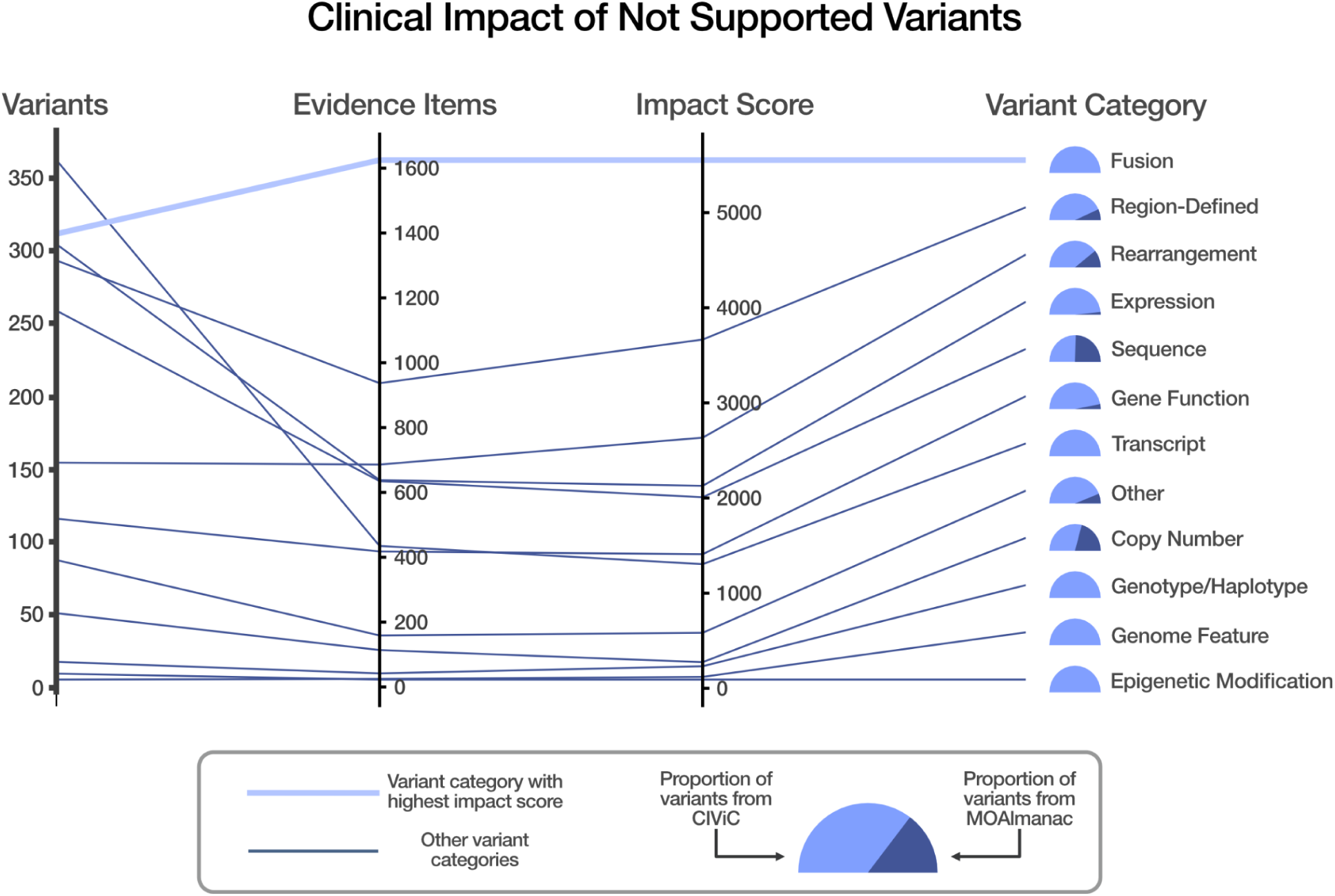
The Clinical Impact of Not Supported Variants. The above plot shows each category of variants outside the current scope of the Variation Normalizer (i.e., not supported variants) along with (from left to right) the number of variants in CIViC and MOAlmanac, number of evidence items, and the total impact score for each of these categories. The light blue line highlights the fusion category, which is the category with the highest impact score. Next to each category title is a fuel gauge which represents the ratio of variants from CIViC (in light blue) vs MOAlmanac (in dark blue). For example, all fusion variants are derived from CIViC, whereas 49% of the sequence variants are derived from MOAlmanac.

We identified “fusion variants” as the category of currently-unsupported variants (n=312) with the highest potential clinical impact, with a total impact score of 5,552 across 1,625 evidence items. Following fusions, region-defined categorical variants (n = 294) such as “MET Exon 14 Skipping Mutations” have 937 associated evidence items, with a total impact score of 3665.5, and other rearrangement variants (n = 157) have associated 686 evidence items, with a total impact score of 2,634.5.

## Discussion

The clinical variation evidence landscape today is starkly different across germline disorders and somatic cancer contexts, in both quantity of available evidence and the complexity of the variants associated with that evidence. This contrast is reflected in the degree to which modern variant representation frameworks are able to associate knowledge across these two clinical domains. We demonstrate that while ClinVar (the predominant germline variant classification resource) is overwhelmingly composed of small genomic variants and CNVs, somatic cancer knowledgebases have a much more diverse representation of variant concepts that are not well-supported by current variant representation standards.

Among the variant types commonly seen across all clinical knowledgebases, normalization methods supporting short variant matching (substitutions and indels) were straightforward to implement and had very high normalization rates compared to other forms of variation. These were the most prevalent variant types across community resources, and this work was enabled by the widespread availability of established community resources for variant liftover (we used the St. Jude chainfile software; see **methods**) and preferred transcript selection (from the MANE project^24^) supporting normalization methods. Notably, the greatest challenges for this class of variation stemmed from the ambiguities surrounding ranged indels in transcript-defined variants lacking associated genomic coordinates, similar to challenges observed in some MAVE datasets^26^. Community conventions for representing such events are needed to inform normalized representation of these events for variant search.

The second most common class of variants were CNVs, where we observed a notable improvement in matching real-world CNV calls to the ClinVar resource by measuring coding sequence overlap over exact base-pair alignment. While a useful proof of principle for a generalized CNV matching strategy, we recognize additional work is needed to refine our match criteria based on other biologically relevant criteria beyond simple CDS overlap. These additional criteria will also be needed for the matching and aggregation of other forms of structural variation (e.g. translocations and inversions), which were not evaluated in this study. Where possible, we will add support for CNV features necessary for technical interpretation^27^, while continuing to work with the broader clinical and informatics communities to identify additional clinically relevant features. Broader community efforts to define these criteria are needed.

Our assessment of remaining forms of variation in somatic cancer knowledgebases emphasized the need for standardized approaches to representing and searching fusion variants. This reflects the established clinical significance of gene fusions as driver events for cancer development^,28,29,30^. Recent advances in community defined information models^31^ and data standards are anticipated to help with this challenge. While there were fewer region-defined and rearrangement variants than fusions across the evaluated somatic cancer knowledgebases, evidence items associated with these classes of variants also demonstrated strong potential clinical impact based on our variant type impact assessment.

Region-defined variants have been challenging to normalize due to their categorical nature. For example, region-defined variants include “*BRAF* mutation” and “*POLE* mutation.” These region-defined variants are broad categorizations that are broadly interpreted to include any non-silent mutation in the respective gene. This concept has historically lacked a common community standard that precisely models the variation represented by such statements. Recent community efforts in defining categorical variant representation has resulted in the GA4GH Categorical Variation Representation Specification (Cat-VRS; cat-vrs.ga4gh.org), a recently released, computable schema to address the challenges surrounding the modeling and normalization of categorical variation. Strategies and tools for matching to this class of variant are currently under development by that community.

It is anticipated that advancements in the genome reference space, such as the Telomere to Telomere (T2T) project^32,33^ and the Human Pangenome^34^, will eventually need to be considered in the representation and search of clinical genomic knowledge resources. These novel reference assemblies are advancing our understanding of the human genome and have the potential to identify additional clinically-relevant variants^35^, and it is anticipated by the Human Pangenome Reference Consortium that there will be an increasingly broad set of clinical applications emerging from the use of pangenome assemblies^34^. Despite these advancements, it is also worth noting that clinical laboratories have been slow to uptake even the GRCh38 reference assembly (first released in 2013), citing a lack of resources and perceived benefit to switching^36^. We anticipate a similar lag in the clinical adoption curve for these relatively recent developments in genome references, though this may be ameliorated through the active participation of these emerging genome reference providers in the clinical variant representation community. For example, the recent development of the GA4GH RefGet Sequence Collections^37^ standard is intended to standardize the use of these newer reference genomes among a landscape of other community genome assemblies.

In conclusion, we have found that systematic normalization of variant representation across knowledge resources is both feasible and essential to overcoming the long-standing interpretation bottleneck in clinical genomics. By harmonizing real-world data with community knowledgebases through the Variation Normalizer, we provide a practical model for unifying genomic evidence across heterogeneous systems, genome builds, and nomenclatures. This work underscores how open, standards-based frameworks—particularly those emerging from GA4GH and related initiatives—can enable reproducible and automated variant interpretation within clinical workflows. Such normalization strategies will be increasingly vital to maintaining interoperability in an increasingly diverse genomic knowledge ecosystem. Continued collaboration between tool developers, standards organizations, and clinical laboratories will be necessary to extend these methods to more complex forms of variation to narrow the remaining gaps in scalable variant interpretation.

## Methods

### Variation Normalizer

The Variation Normalizer is an open-source Python package and REST API available at https://zenodo.org/records/16112010. The tool translates human-readable variation descriptions to normalized VRS representations. It supports free-text, VCF-like, and HGVS expressions^38^ for protein, coding DNA, or genomic DNA reference sequences (as applicable). The REST API has several endpoints. The default variant endpoints are *to_vrs* and *normalize*. The CNV endpoints are *hgvs_to_copy_number_count*, *hgvs_to_copy_number_change*, *parsed_to_cn_var*, *parsed_to_cx_var*, and *amplification_to_cx_var*. Endpoints from other services, such as VRS-Python^39^ and Cool-Seq-Tool^23^, are also exposed for added functionality.

The Variation Normalizer processes and normalizes queries in four discrete steps: tokenization, classification, validation, and translation. During *tokenization*, query strings are split on whitespace. Each substring is provided as input to each tokenizer. The tokenizers perform regular expression searches and check other conditions to determine if the string is a match for its respective token type. The Gene Normalizer^40^ is used during this step to help identify gene tokens. The *tokenize* method returns an ordered list of tokens.

During *classification*, the ordered list of tokens is passed to each classifier. Every classifier has an *exact_match_candidates* method which returns a list of lists containing the ordered tokens needed for that classification. The classification step only performs exact matching and will only return a single classification. The resulting classification is used during the validation process.

During *validation*, Ensembl and RefSeq data harvested from Cool-Seq-Tool is used to retrieve a list of compatible accessions for the given classification. If the classification’s nomenclature is *free_text*, typically more than one accession is returned for the associated gene. For *gnomad_vcf* nomenclature, the related GRCh37 and GRCh38 accessions will be returned. For *hgvs* nomenclature, the provided accession will be used. Several checks are made for each accession identified. The Biocommons SeqRepo^41^ is used to validate accessions and check that the expected reference sequence matches the actual sequence for the classification’s start and end positions. The Gene Normalizer is used again to help determine if the classification’s positions exist on the related gene. HGVS nomenclature syntax is validated by ensuring positions are unique and listed from 5’ to 3’. For a protein classification, amino acid codes are validated using Biocommons Bioutils^42^. The validation step will return a list of valid results and a list of invalid results.

During *translation*, the valid results are passed as input to each translator. During this step, each valid result is translated to a VRS variation. The *normalize* method uses Cool-Seq-Tool to get the MANE representation through its transcript priority algorithm (**Supplemental Figure 3**). Another Genome Conversion Tool (agct)^43^ is a drop-in replacement for PyLiftover^44^ built on top of the St. Jude chainfile software (https://github.com/stjude-rust-labs/chainfile), which we use to map the starting annotation to the preferred GRCh38 assembly. Cool-Seq-Tool then maps to the MANE representation using the latest MANE summary data and the Biocommons Universal Transcript Archive^45^ database, which stores transcript alignment data. Cool-Seq-Tool will then map back to the starting annotation layer. If a MANE representation is not found, the longest compatible remaining transcript representation will be returned. Other methods, such as *to_vrs*, will not perform any liftover. For all methods (*normalize*, *to_vrs*, etc.), VRS-Python models are used to create the VRS variation and fully-justified allele normalization is applied. Then, globally consistent and unique computed identifiers are generated for the *VRS variation*. Finally, each translator will return a list of translation results containing information such as the *VRS variation* and status of the accession in the *VRS variation*. In the *to_vrs* method, all translation results will be returned. When using the *normalize* method, only one translation result will be returned based on prioritized accession status (in order from most to least priority: “*mane_select*”, “*mane_plus_clinical*”, “*longest_compatible_remaining*”, “*grch38*”, or “*na*”).

The Variation Normalizer has internal rules when multiple translation results are “tied” in the *normalize* method. A tie happens when the most prioritized accession status has numerous, different translation results, each with the same count. If there’s more than one preferred translation, the translation results will be sorted by the original accession used to get that translation result. If the original accession matches the accession found in the *VRS variation*, that result will be returned. If there are no matches, the first translation result in the sorted list will be returned.

The normalize method also has a hgvs_dup_del_mode parameter. HGVS Dup Del Mode is an algorithm to help interpret deletions and duplications that are represented as HGVS expressions. There are several modes: default, copy_number_count, copy_number_change, repeated_seq_expr, and literal_seq_expr (**Supplemental Figure 4**).

The Variation Normalizer has several methods for representing VRS Copy Number Variation. There are methods for using HGVS expressions to return a *VRS Copy Number Change* (*hgvs_to_copy_number_change*) or a *VRS Copy Number Count* (*hgvs_to_copy_number_count*). These methods re-use the *tokenize*, *classify*, *validate*, and *translate* methods described above. There are also methods for taking parsed parameters and returning a *VRS Copy Number Change* (*parsed_to_cx_var*) or a *VRS Copy Number Count* (*parsed_to_cn_var*). These methods construct Copy Number models from the input parameters. The *amplification_to_cx_var* method takes a gene or location data and returns a VRS Copy Number Change.

### GENIE Analysis

The *data_mutations_extended.txt* dataset^46^ from AACR project GENIE^17^ 18.0-public release was downloaded for the purpose of evaluating patient variant matching to different genomic knowledgebases. The following columns were selected from the dataset for downstream analysis: *Hugo_Symbol*, *NCBI_Build*, *Chromosome*, *Start_Position*, *End_Position*, *Tumor_Sample_Barcode*, *Reference_Allele*, *Tumor_Seq_Allele2*, and *HGVSp_short*. Two new columns were then added to the dataset: *free_text_p_short* (a protein variant in the format *{Hugo_Symbol} {HGVSp_short}*) and *coordinates* (a genomic variant in the VCF-like format *{Chromosome}-{Start_Position}-{Reference_Allele}-{Alternate_Allele}*) (“GENIE pre-variant analysis notebook” on GitHub; see **Data Availability**). Records that were missing requisite values for these columns were excluded from the analysis, as were rows with duplicate expressions. The unique protein and genomic expressions were each provided as input to the Variation Normalizer’s *normalize* method. If the GRCh37 genomic expression failed to normalize (e.g. due to failed liftover to GRCh38), the expression was then passed to VRS-Python’s *translate_from* method, which does not perform liftover operations. 86.25% (823,070/954,230) of protein expressions and 99.88% (962,718/963,850) of genomic expressions were able to normalize.

Following the normalization procedure, matching to the CIViC, MOAlmanac, and ClinVar knowledgebases was attempted at both the variant and patient level. First, dataframes of normalized variants from CIViC, MOAlmanac, and ClinVar (see below) were ingested and joined with two dataframes of normalized protein and genomic GENIE variants, as well as a combined dataframe (containing duplicate expressions) from the pre-variant-analysis notebook (genie_search_analysis notebook on GitHub; see **Data Availability**). Next, a series of set intersection operations were performed to determine the shared VRS identifiers between the normalized GENIE variants and the normalized variants from CIViC, MOAlmanac, and ClinVar. This resulted in the following lists of shared variants: GENIE protein and CIViC, GENIE genomic and CIViC, GENIE protein and MOAlmanac, and GENIE genomic and ClinVar.

After deriving lists of shared variants between GENIE and the knowledgebases, we attempted to perform matching at the patient sample level, where the presence of at least one shared variant indicated a match. To conduct this analysis, two columns were added to the GENIE variants dataset: a *vrs_id_genomic* column, reporting the VRS identifier for genomic variants; and a *vrs_id_protein* column, reporting the VRS identifier for protein variants. If normalization was not successful for a variant, its VRS identifier was set to *N/A*. An additional four match columns were added to the dataset, describing if a VRS identifier was found in a knowledgebase (e.g., *in_civic_protein* and *in_clinvar*). Finally, two dictionaries were created, one at the protein level and one at the genomic level, where each key was a patient sample identifier and its associated data was that sample’s list of variants. To count the number of patient sample matches, a set intersection operation was performed between each sample list and the corresponding list of shared variants; if the length of the intersected list was >= 1, a match was recorded. This process was completed four times, once for each match column that was added to the GENIE dataset.

After evaluating matching at the patient sample level, we then determined the average number of protein and genomic variants per sample that were contained in each knowledgebase. First, the average success of variant normalization per patient was calculated by dividing the average number of total variants per patient by the average number of normalized variants per sample. Then, the GENIE dataset was filtered to variants present in each knowledgebase, and also by the variant type (protein or genomic). The average number of variants per patient sample was then calculated under for each subset. Lastly, these values were then added to three tables (see **Supplemental Table 4**) to allow for data visualization and interpretation.

### Nationwide Children’s Hospital (NCH) Microarray CNV Cohort

Laboratory test results were obtained for 22,593 microarrays ordered at NCH between October 3, 2006 and May 29, 2023. Regular expressions were used to parse ISCN nomenclature strings for CNVs from the resulting free-text reports, and to parse the chromosome, start position, stop position, and copy number range of each CNV (**Supplemental Figure 5**). For CNVs with an exact copy number, both the minimum and maximum of the range were considered to be equal to the exact copy number.

Of the unique ISCN strings extracted, any that were malformed, represented regions of homozygosity, or were mapped to NCBI36 were excluded. Each of the distinct variants remaining was normalized using the Variation Normalizer’s *parsed_to_cn_var* endpoint. Normalized variants were used in subsequent CNV analyses (**Supplemental Figure 1**)

### Matching Microarray CNVs to ClinVar CNVs

We sought to measure how many of the 8,718 normalized NCH CNVs could be matched to variants among the 71,730 normalized ClinVar CNVs. The first challenge we encountered was the reality that many CNVs were assayed by probes with limited granularity, making the start and stop positions knowable only within a margin of error. This phenomenon was observable in HGVS expressions of the form *NC_######.#:g.(<outer_start>_<inner_start>)_(<inner_end>_<outer_end>)*, representing ranges of possible start and end positions for a CNV. We chose to represent any CNV by its most specific known positions, preferring innermost start and end coordinates in cases where exact start or stop positions were not known.

The primary difficulty in matching these large variants lies in the existence of CNVs which occupy similar but not identical regions of the genome. To rigorously quantify suitable notions of similarity, we calculated two different Jaccard scores to measure the similarity between a given pair of CNVs (**Figure 2C**)

For a given NCH query variant *Q* and candidate ClinVar variant *C*, the base pair (BP) score was calculated as 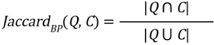. This amounts to the proportion of the length of sequence overlapped by both variants out of the total length of sequence spanned by either variant. This score function takes values in the range [0, 1]. If *Jaccard*_*BP*_(*Q*, *C*) = 0 then *Q* and *C* have no base pairs in common; if they span the exact same base pair range, then *Jaccard*_*BP*_(*Q*, *C*) = 1.

This is a straightforward metric for measuring positional similarity of CNVs, but such a scoring scheme has shortcomings. For instance, it may give a high similarity score to a pair of variants which overlap on long regions of noncoding DNA but which share very little common protein coding DNA, or it may punish pairs of variants with similar protein consequences but which also contain large stretches of differing noncoding DNA. To account for this, we also measured the similarity score based only on the coding regions between pairs of CNVs and termed this the CDS Jaccard score.

To calculate the CDS Jaccard score, we calculated the regions of every normalized CNV from ClinVar or the NCH data with coding sequences (CDS) by providing the *feature_overlap* endpoint with normalized CNV start and stop positions (**Supplemental Figure 2**). This linked each CNV to the locations on which it overlapped any protein coding sequences as recorded in the MANE RefSeq GFF file (https://ftp.ncbi.nlm.nih.gov/refseq/MANE/MANE_human/release_1.4/MANE.GRCh38.v1.4.refseq_genomic.gff.gz).

The CDS Jaccard score is formulated as the ratio of the combined size (in number of base pairs) in coding regions encompassed by both the query and the candidate variant to the size of coding regions contained in either. That is, letting *Q*_*CDS*_ and *C*_*CDS*_ be the subsets of base pairs in coding regions overlapped by *Q* and *C*, respectively, 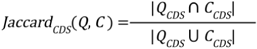 Similarly to *Jaccard*_*BP*_, *Jaccard*_*CDS*_ takes values in the range [0, 1]. Notably, a pair of variants must have nonzero BP similarity (*Jaccard*_*BP*_(*Q*, *C*) > 0)) in order to have nonzero CDS similarity. However, it’s possible to have variants that overlap entirely on noncoding regions so that *Jaccard*_*BP*_(*Q*, *C*) > 0 but *Jaccard*_*CDS*_(*Q*, *C*) = 0.

In addition to the ambiguity of similarity of genomic location of CNVs, we accounted for agreement or disagreement of copy number by considering three different matching policies of increasing strictness: *Ignore CN* considered only the BP or CDS overlap of two variants without accounting for the copy number state of either variant. *No CN Disagreement* allowed two CNVs to be considered a match as long as their copy number ranges overlapped *or* if copy number information was unavailable for one or both of the variants; for example, under this policy, a variant with copy number range [3, 4] could match with a variant with copy number range [2, 3] or with no known copy number, but not with a variant with copy number 1. *CN Agreement* required that both variants have known copy number ranges and that they overlap; for example, under this policy, a variant with copy number range [3, 4] could match with a variant whose copy number is 3, 4, or whose copy number range is any interval which intersects [3, 4].

For each query variant in the set of normalized NCH CNVs and each candidate variant in the set of normalized ClinVar CNVs, both BP and CDS Jaccard similarity scores were measured (**Figure 2C**) and the number of NCH variants with at least one match was calculated for similarity thresholds ranging from 0 to 1 (**Figure 2D**).

### Variant Category Creation

In order to assess the Variation Normalizer’s performance and highlight opportunities for future development, we generated a coarse ontology of variant types in CIViC and split the variant tokens from the dataset into categories. We chose CIViC as the basis for this analysis for two reasons. First, CIViC is a community-sourced knowledgebase, so the variant entries extracted from the literature and submitted to CIViC cover a broad range of variant types and reflect the submitter’s understanding of categories related to those variants. Second, CIViC entries are curated by both CIViC editors and Somatic Cancer Variant Curation Expert Panels (SC-VCEPs), who verify the categorical intuition of the original submitters and evaluate the strength of the evidence attached to each variant. We therefore used CIViC as a reasonable approximation of the underlying understanding of variant categories in the broader research community.

We generated a typology of variant categories by recursively grouping variants by degree of similarity in terms of the properties of each variant in CIViC. We first constructed an ontology log^47,48^ to model categorical variants in CIViC. Because the domain of the ontology log is the category of SET, the objects are sets of variants and the arrows are morphisms between those sets. The individual members of each set share certain core properties, and also may or may not share other optional attributes. For instance, all variants in the set of fusions entail a change in location. Some fusions may also affect protein expression, but this property is not required to conclude whether or not a given variant is a member of the set of fusions.

We performed a distributional analysis^49^ on the resulting ontology log to collapse variant sets by similarity in their core properties. This process merges individual variant categories into broader supercategories in a recursive bottom-up fashion. For example, the sets of in-frame deletion variants and out-of-frame deletion variants only differ by one core property, and can be collapsed into their mutual superset, the set of deletions. Deletions and insertions can be similarly collapsed into their mutual superset of sequence variants.

The resulting typology of variants and distinguished variant categories (*Sequence, Genotype/Haplotype, Fusion, Rearrangement, Epigenetic Modification, Copy Number, Expression, Gene Function, Region-Defined, Genome Feature, Transcript,* and *Other variants*) along with seven core properties are reflected in **Table 1**. The first five properties are related to the ways in which an assayed variant can differ from the reference genome: a change in sequence, a change in location, a change in reading frame, a change in quantity, or else a change in function. The sixth property relates to the biological level at which the variant was assayed. This allowed us to meaningfully distinguish, for instance, between genomic sequence variants, *Sequence Variants*, which are overwhelmingly supported by the Variation Normalizer, from systemic-level variants, *Genotype Variants*, such as Loss of Heterozygosity variants, which are not in scope for the Variation Normalizer. A similar contrast appeared for Copy Number Variants which are supported, versus Epigenetic Modification variants which are not.

The final property concerns a variant being located at a particular locus of interest. Variants in all the categories shown in **Table 1** admit region specificity as at least an optional property. Any variant for which only the region property is specified is collapsed into the category of *Region-Defined Variants*. Examples of categorical variants that fall into this set include “*EGFR* mutations”, “3’ UTR Mutations”, and “*TP53* Mutations.” Finally, the category of *Other Variants* in **Table 1** contains any variants that are not identifiable as being a member of one of the other categories, often due to the entry being incomplete or making use of nonstandard notation conventions. One example of variants in this category is the category of *Rare Mutations*, because no other information (other than relative frequency) is provided about these variants in order to admit them into a different category. In sum, we found that most categorical variants can be usefully distinguished according to the way in which it differs from the reference genome, and biological level. *Region-Defined Variants* are those variants where only the region is known but nothing else, so it cannot be classified with confidence, and *Other Variants* are those which may have any combination of properties, but lack sufficient detail as to make further classification possible.

### CIViC Variant Normalization

CIViCpy^50^ was used to get CIViC data on July 17th, 2025. Variants and their associated entities matching *accepted* and *submitted* statuses were retained which led to a total 3,845 variants. Each CIViC variant was categorized as a protein or a genomic base representation. For all small variants, a CIViC variant name containing a *“c.”* indicates a contextualized genomic variant whereas all others were considered protein variants. For genomic variants, the HGVS genomic expression associated with the variant was used as the base representation. If no HGVS genomic expression was found, the representative variant coordinates were transformed to create a VCF-like format (*{chromosome}-{start}-{reference_bases}-{variant_bases}*). At this time, CIViC only supports the GRCh37 assembly. For protein variants, the base representation used was *{gene name} {variant name}*.

Variant categories were created to bin the variants that are known to be out of scope for the Variation Normalizer: *expression*, *epigenetic*, *fusion*, *sequence*, *gene function*, *rearrangement*, *copy number*, *genotype*, *region-defined*, *transcript*, and *other*. *Transcript variants* are CIViC genomic variants where no HGVS or representative coordinate was found. A mapping was created for each unsupported category and variant names that belong in the category. Base representations of variants were checked via this mapping and through the use of regular expressions to determine if the CIViC variant is out of scope of the Variation Normalizer and if so, determine which unsupported variant category it belongs within. Such variants were not run through the Variation Normalizer.

### Molecular Oncology Almanac Feature Normalization

The MOAlmanac features were fetched via the REST API feature endpoint, https://moalmanac.org/api/features, on July 17th, 2025. The total number of features (variants) were 452. A query was created for each MOAlmanac feature which depended on the feature type (notebook on GitHub; see **Data Availability**). Variant categories were created to bin the features that are known to not be supported in the Variation Normalizer: *expression*, *epigenetic*, *fusion*, *sequence*, *gene function*, *rearrangement*, *copy number*, *genotype*, *region-defined*, *transcript*, and *other*. A mapping was created from each MOAlmanac feature type to the categories of variants out of scope by the Variation Normalizer. Somatic and germline feature types were not included in this mapping and required the *variant_annotation* and *exon* fields to determine the category of unsupported variants. Queries were checked via this mapping and additional checks were performed to determine if the MOAlmanac feature is not supported by the Variation Normalizer and if so, determine which unsupported variant category it belongs to. Such features were not run through the Variation Normalizer.

### ClinVar Variant Normalization

#### Categorization

The Variation Normalizer attempted to generate a VRS representation of each ClinVar variant. The vast majority of variants in ClinVar were represented by a Canonical SPDI which is based on build GRCh38 and is constrained to variants of length up to 50bp. The next significant tranche are identifiable by either their build GRCh38, GRCh37 or NCBI36 genomic HGVS expression in order of priority. Next, remaining variants are identified by their build GRCh38, GRCh37 or NCBI36 genomic location data. if available. Any remaining variants were not attempted to be VRSified nor normalized. CNVs were identified by being either typed by ClinVar as copy number gain/loss or being deletions or duplications of length greater than 1000bp. Some CNVs in ClinVar have absolute copy numbers specified in the data which were targeted as CopyNumberCount VRS classes, while the others were targeted as CopyNumberChange variants depending on whether they were gains/duplications or losses/deletions.

The normalizer directly utilized the Canonical SPDI or HGVS representations if available to generate a VRS representation of variants that were not CNVs. HGVS expressions and either absolute copy number or copy change type gain/loss was provided to the normalizer for all CNVs.

Next, we processed the variant description to categorize the ClinVar variants into one of the following bins, listed in order of precedence: *Allele* (variants normalized to a Canonical SPDI format); *Genotype and/or Haplotype variants*; *Unsupported HGVS expressions*; *CopyNumberCount* (Copy Number variants with an absolute copy count value); *Copy Number variant with a min / max count range*; *CopyNumberChange* (CNV loss/gain, and Deletion or Duplication variant types greater than 50 nucleotides); *Remaining valid HGVS* variants; and *No HGVS and location data*.

#### Normalization

For each of the binned groups above, the preferred format and type were used to determine which Variation Normalizer REST API endpoint to call: *Allele* - Canonical SPDI and remaining HGVS variants used the *translate_from* endpoint, *CopyNumberCount* used the *hgvs_to_copy_number_count* endpoint, and *CopyNumberChange* used the *hgvs_to_copy_number_change* endpoint.

For *CopyNumberChange*, we applied one of two VRS maintained Experimental Factor Ontology^51^ (EFO) relative copy number variation labels for *copy number gain* and *copy number loss* based on the ClinVar variant type. For ClinVar variation types of *copy number gain* or *Duplication*, the copy number gain label “gain” was used. Finally, for types of *copy number loss* or *Deletion*, the copy number loss label “loss” was used.

### Impact Analysis

Variants were sourced from CIViC and MOAlmanac databases. CIViC evidence items and MOAlmanac assertions were also extracted; we will refer to both as evidence items henceforth. CIViC evidence items can have a *submitted, accepted*, or *rejected* status. Only *accepted* and *submitted* CIViC evidence items were used in the analysis. Some evidence items can be applicable to multiple variants. Next, evidence items were classified based on the associated variation normalization status. Three groups were created: evidence items with variants that were normalized, evidence items associated with variants that failed normalization, and evidence items with variants that are not supported by the Variation Normalizer (not in the current scope of the Variation Normalizer). Before conducting the analysis, duplicate evidence items were removed. Additionally, among the variants that were not supported, evidence items were further segregated based on the subcategory of variants to which they were linked (see CIViC Variant Normalization & Molecular Oncology Almanac Feature Normalization). Duplicates were eliminated from each category before proceeding with the analysis.

To calculate an impact score for each variant, we used the MOAlmanac-provided predictive implication and the CIViC-provided evidence level to create a scoring system based on the AMP/ASCO/CAP guidelines^52^. In the case of MOAlmanac, each evidence item had an assigned predictive implication, which was converted into a numerical score following a specific framework: FDA Approved (10), Guideline (10), Clinical evidence (5), Clinical trial (5), Preclinical (1), and Inferential (0.5). Similarly, for CIViC, evidence items were assigned a numerical score based on their evidence level: A (10), B (5), C (3), D (1), and E (0.5). Each variant impact score was calculated by summing the individual evidence numerical scores. Subsequently, the category score was determined by summing the variant impact scores within each category.

The final step involved combining scores for each category from both CIViC and MOAlmanac. Two distinct analyses were conducted: one incorporating both *accepted* and *submitted* evidence items from CIViC along with MOAlmanac evidence items (see results), and another considering only *accepted* evidence items from CIViC in conjunction with MOAlmanac evidence items (**Supplemental Figure 6**). This did not affect the order of the top two most impactful categories.

## Acknowledgements

We acknowledge Evan Christensen (University of Utah Department of Biomedical Informatics), for his role in reviewing the manuscript. We acknowledge Karen Tsuchiya, Katie Schieffer, and Catherine Cottrell (Institute for Genomic Medicine at Nationwide Children’s Hospital) for their contributions to the CNV matching strategy.

The authors would like to acknowledge the American Association for Cancer Research and its financial and material support in the development of the AACR Project GENIE registry, as well as members of the consortium for their commitment to data sharing. Interpretations are the responsibility of study authors.

## Data Availability

Details about this analysis and how to reproduce our results can be found at https://github.com/GenomicMedLab/variation-normalizer-manuscript. The notebooks used [v0.15.0](https://github.com/cancervariants/variation-normalization/releases/tag/0.15.0) of the Variation Normalizer. This repository contains Jupyter notebooks that demonstrate how to utilize the Variation Normalizer and match evidence from knowledgebases to assayed variant data.The *download_s3_files* notebook provides a way to programmatically download the files from a public s3 bucket. All content is freely available for academic research.

GENIE data from release 18.0 was not included because it requires a Synapse account for access. An account can be created at: https://www.synapse.org/#!RegisterAccount:0.

## Code Availability

The Variation Normalizer source code is available at https://github.com/cancervariants/variation-normalization. The Jupyter analysis notebooks are available at https://github.com/GenomicMedLab/variation-normalizer-manuscript. Both are freely available for reuse under the MIT license.

## Notes

### Competing Interest Statement

The authors have declared no competing interest.

### Funding Statement

This study was funded by the National Human Genome Research Institute of the National Institutes of Health, project R35HG011949.

### Author Declarations

IRB of Nationwide Children's Hospital gave ethical approval for this work under NLP for Genomic Medicine study (STUDY00000276).

### Summary of Updates

Corrected author list typos from original submission

