## Supplemental Figures and Tables for "The Clinical Genomic Variation Landscape"

#### Criteria for Selecting Variants from NCH Microarray Reports

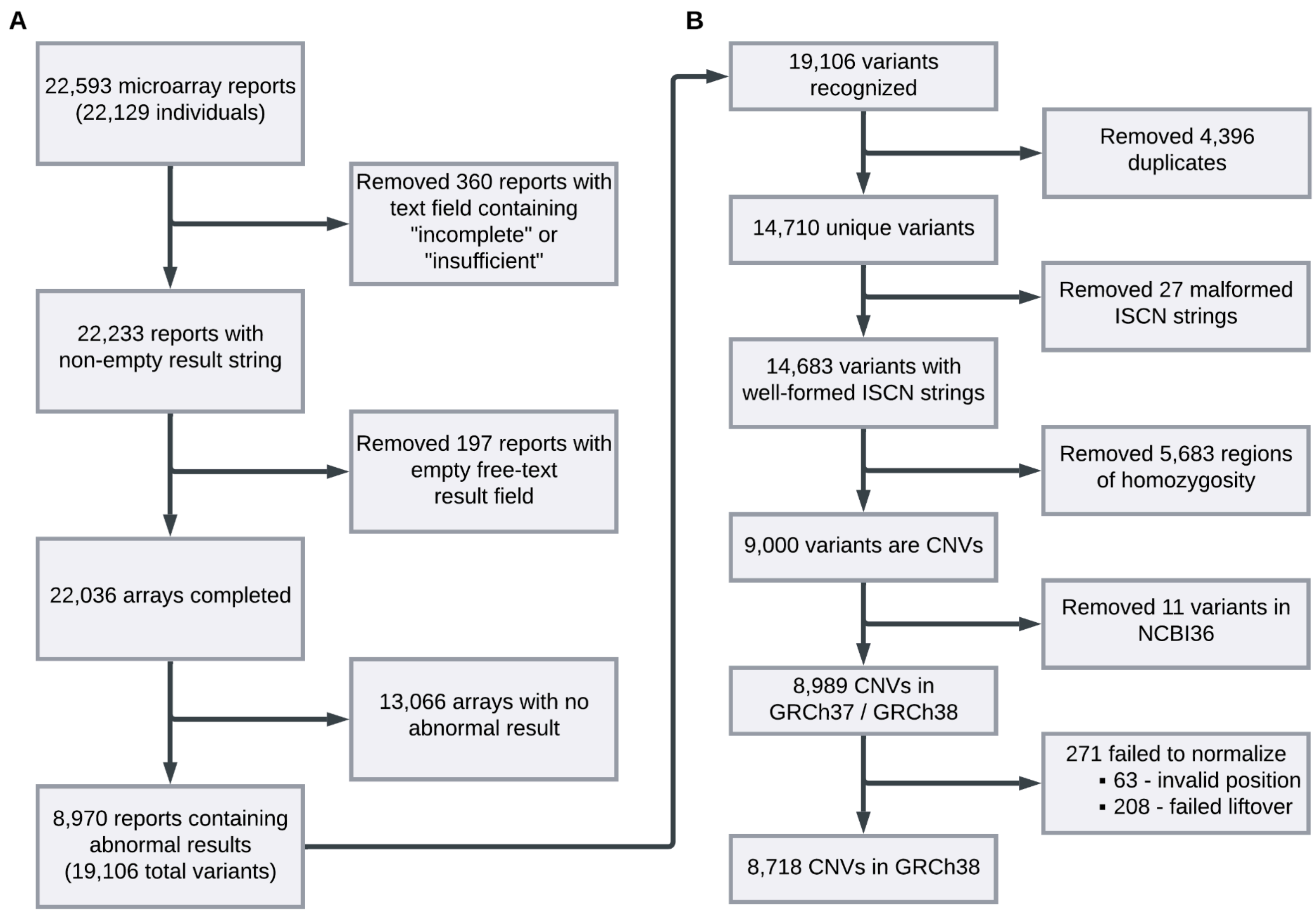


**Supplemental Figure 1: Criteria for Selecting Variants from NCH Microarray Reports.**

(a) A depiction of the criteria that were used to extract CNV ISCN strings from free-text reports. Reports were dropped from the analysis for having features including missing or insufficient text fields and including arrays without abnormal results. (b) The variants extracted from free text, with exclusions and failure modes in the Variation Normalizer. CNV variants were ultimately dropped for reasons including having malformed ISCN strings and being described on the NCBI36 build.

#### NCH and ClinVar CNV Coding Overlap

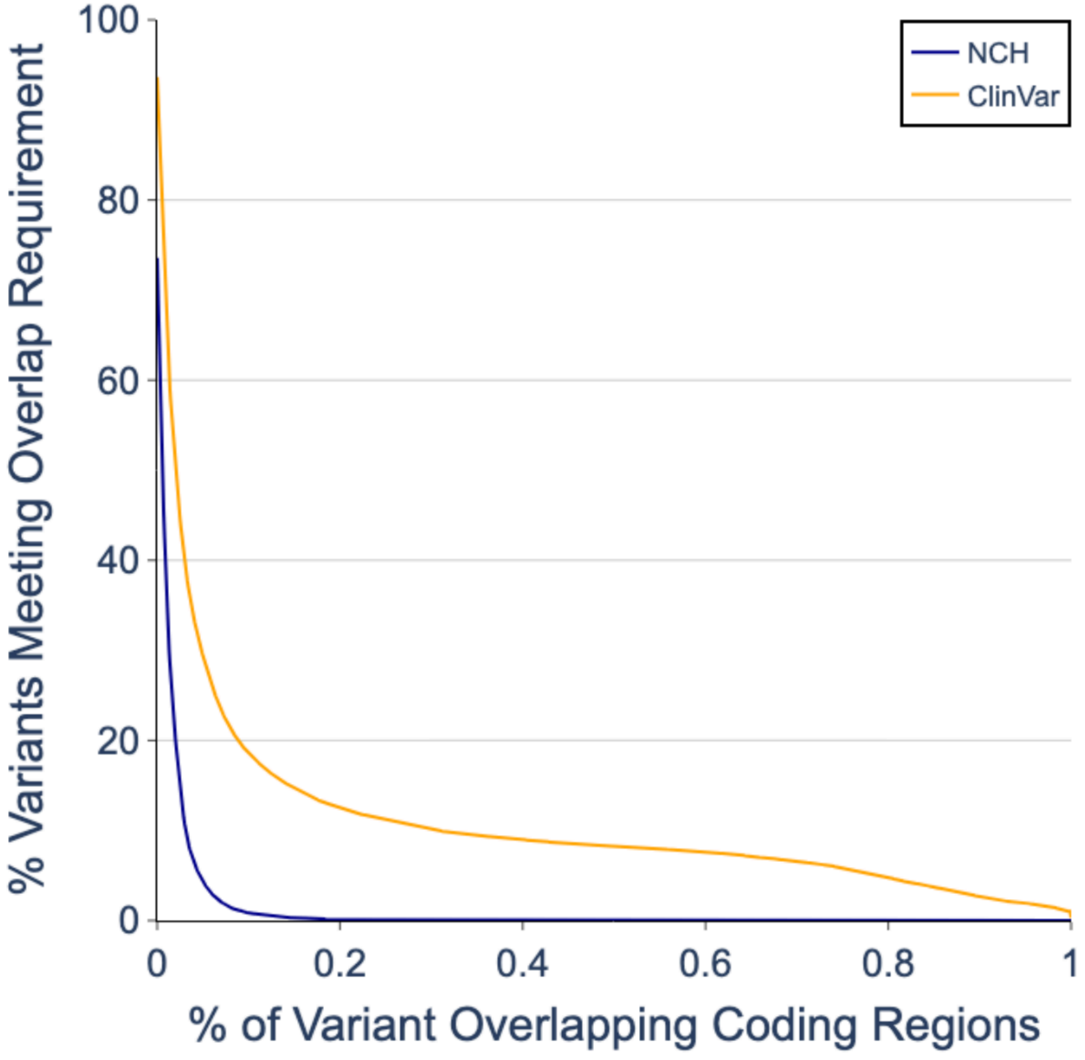


#### Clinical Impact of Not Supported Variants

**Supplemental Figure 2: Overlap of NCH and ClinVar Variants with Coding Regions.** The horizontal axis shows a range of percentage overlap with coding regions and the vertical axis measures the percentage of CNVs meeting the given percentage overlap requirement. NCH percentages are shown in blue and ClinVar percentages are shown in orange.

### Cool-Seq-Tool Transcript Priority Algorithm

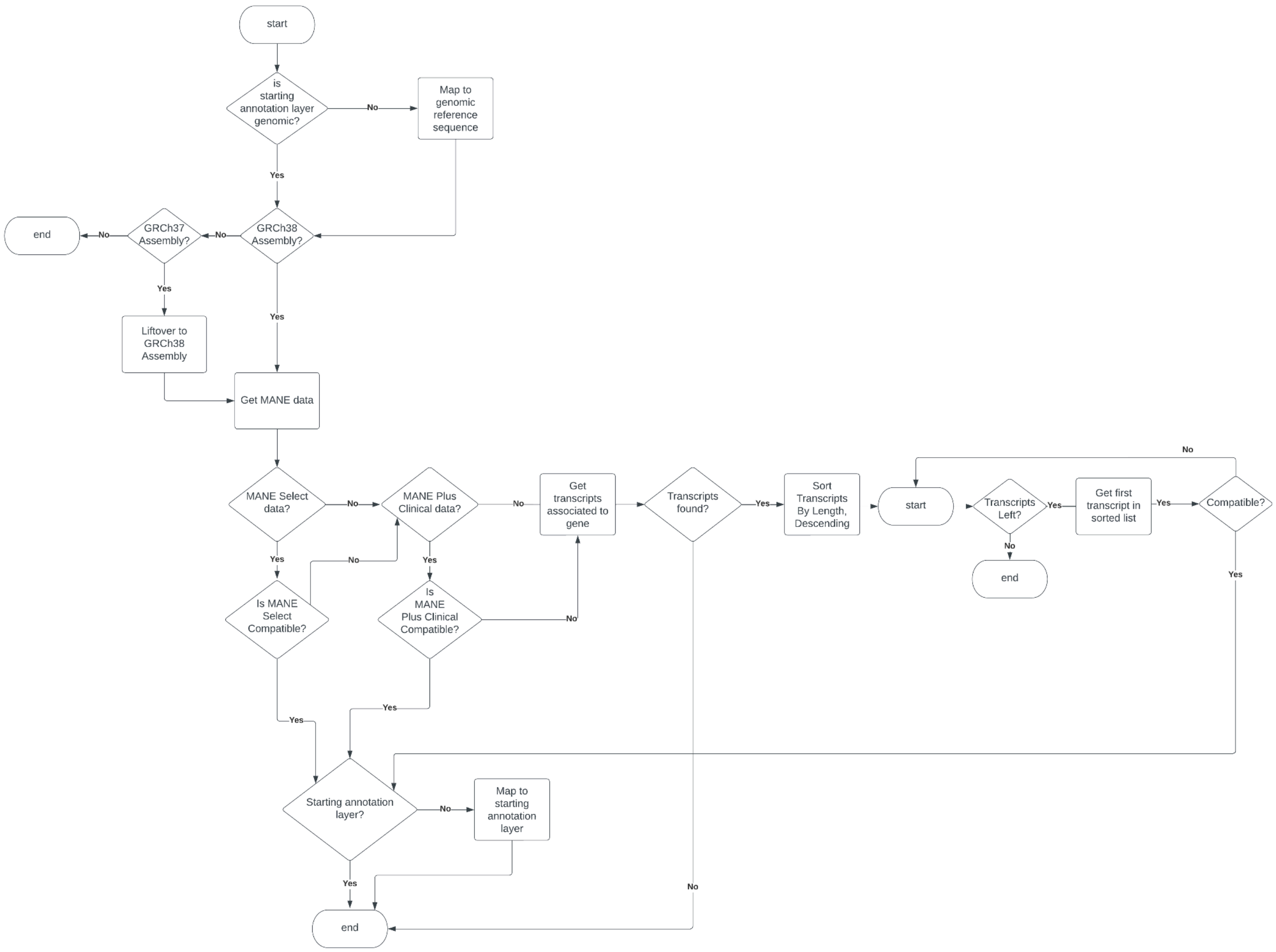


**Supplemental Figure 3: Cool-Seq-Tool Transcript Priority Algorithm.**

The workflow demonstrates the policy for selecting a representative transcript using sequence attributes and MANE annotations.

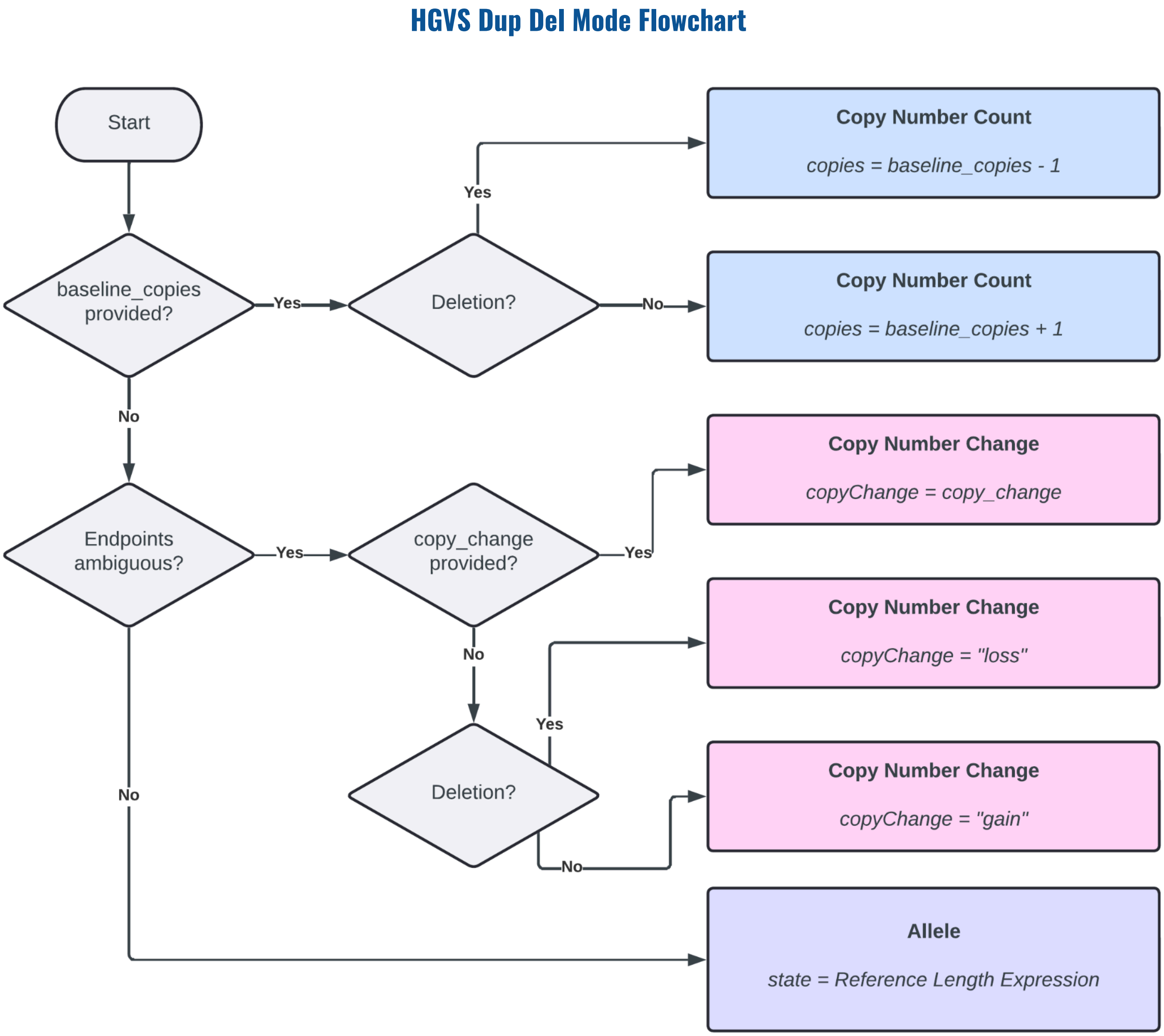


**Supplemental Figure 4: HGVS Dup Del Decision Flowchart.** The decision flowchart for interpreting deletions and duplications that are represented as HGVS expressions. Copy number variant representation can be ambiguous, so we created an algorithm to help us interpret these expressions. Ambiguous endpoints are any uncertain ranges, such as the form (#_#)_(#_#).

#### Regular Expression for Detecting ISCN Nomenclature

```
([XY\d]+[pq][XY\d\/\.pq]+\([\d,\-\_\(\)]+\)?x ?[\d\~\-]+ ?(?:hmz)?)
```

**Chromosome**

**Arm**

**Band/sub-band**

**Start/Stop positions**
*ISCN allows comma-separated decimal numbers
*conventions have separated start and stop both by "-" and "_"

**Copy Numbers**
Nomenclature allows for ambiguous copy number ranges

**Regions of homozygosity**

**Supplemental Figure 5: Regular expression for Detecting ISCN Nomenclature strings.** This regular expression was used with the python re.findall function to extract ISCN strings for CNVs from the text of microarray reports. The boxes below the expression summarize the purpose of each component.

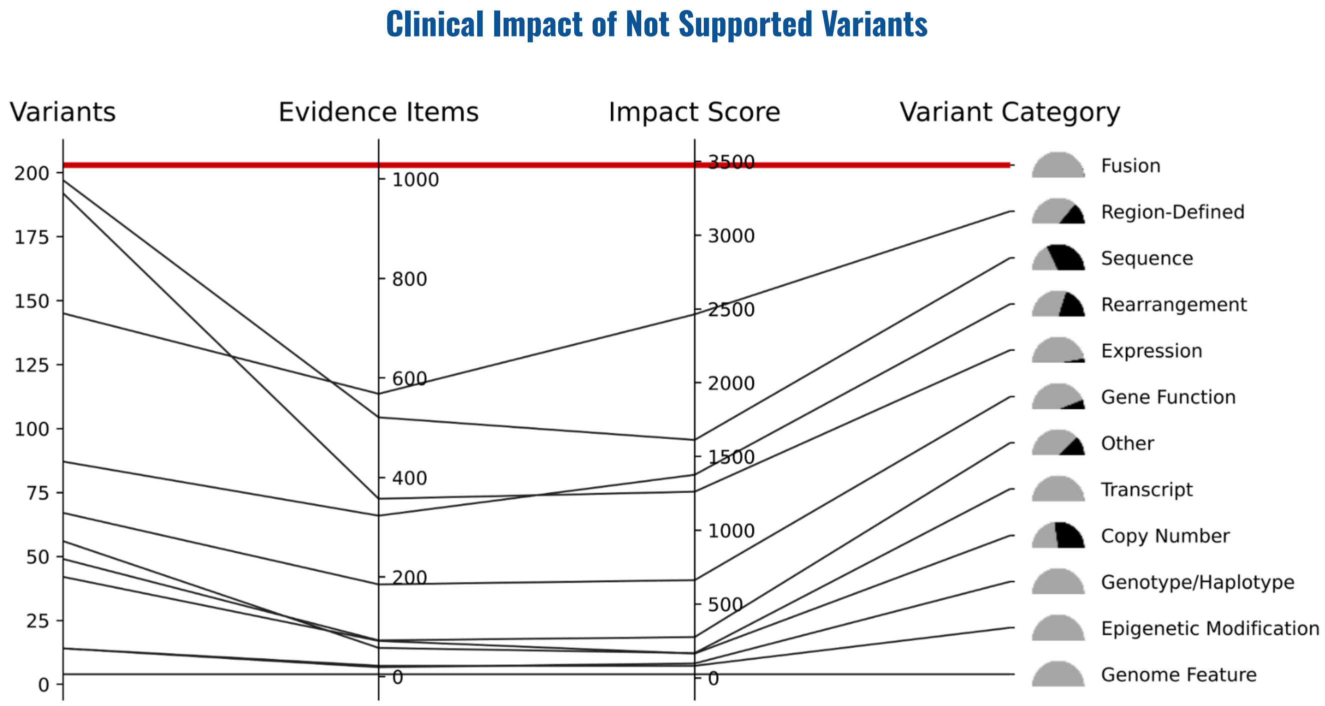


**Supplemental Figure 2: Overlap of NCH and ClinVar Variants with Coding Regions.** The horizontal axis shows a range of percentage overlap with coding regions and the vertical axis measures the percentage of CNVs meeting the given percentage overlap requirement. NCH percentages are shown in blue and ClinVar percentages are shown in orange.

#### GENIE Normalization & Variant Matching between Knowledge Bases

| Table 1A | Protein Representation | Genomic Representation |
|---|---|---|
| Unique GENIE Variants | 954,230 | 963,850 |
| Unique GENIE Variant Queries Supported by the Normalizer | 823,070 | 962,718 |
| Unique Normalized GENIE VRS IDs | 821,461 | 962,504 |
| Unique Normalized Variants in ClinVar | 0 | 183,780 |
| Unique Normalized Variants in Molecular Oncology Almanac | 143 | 0 |
| Unique Normalized Variants in CIViC | 1,146 | 233 |

| Table 1B | Protein Representation | Genomic Representation |
|---|---|---|
| Average Variants per Patient Sample | 13.17 | 12.10 |
| Average Normalized Variants per Patient Sample | 10.79 | 12.08 |
| Average Normalized Variants per Patient Sample in ClinVar | 0 | 5.59 |
| Average Normalized Variants per Patient Sample in Molecular Oncology Almanac | 1.14 | 0 |
| Average Normalized Variants per Patient Sample in CIViC | 1.46 | 1.02 |

**Supplemental Table 1: Variant and patient sample-level matching of AACR Project GENIE data to various genomic knowledgebases.** Table 1A displays the counts of normalized protein and genomic variant representations in GENIE, and the number of normalized variants that are shared with the ClinVar, MOAlmanac, and CIViC knowledgebases is reported. Table 1B examines matching at the patient sample level, and the average number of normalized variants with a match to an examined knowledgebase is shown.

#### CIViC Normalization Performance

##### A. Normalization of Variants in CIViC

| Category | Percent of all CIViC Variants |
|---|---|
| Normalized | 2015 / 3845 (52.41%) |
| Unable to Normalize | 83 / 3845 (2.16%) |
| Not Supported | 1747 / 3845 (45.44%) |

##### B. Not Supported Variants in CIViC

| Category | Percent of all CIViC Variants |
|---|---|
| Sequence | 133 / 3845 (3.46%) |
| Genotype/Haplotype | 22 / 3845 (0.57%) |
| Fusion | 313 / 3845 (8.14%) |
| Rearrangement | 122 / 3845 (3.17%) |
| Epigenetic Modification | 14 / 3845 (0.36%) |
| Copy Number | 32 / 3845 (0.83%) |
| Expression | 294 / 3845 (7.65%) |
| Gene Function | 111 / 3845 (2.89%) |
| Region-Defined | 255 / 3845 (6.63%) |
| Genome Feature | 10 / 3845 (0.26%) |
| Transcript | 362 / 3845 (9.41%) |
| Other | 79 / 3845 (2.05%) |

**Supplemental Table 2: CIViC Normalization Performance.** (A) The number of variants within CIViC that were successfully normalized, supported but not normalized, or not supported. (B) The number of not supported variants in CIViC sorted by category.

### MOAlmanac Normalization Performance

#### A. Normalization of Variants in MOAlmanac

| Category | Percent of all MOAlmanac Features |
|---|---|
| Normalized | 2015 / 3845 (52.41%) |
| Not Supported | 1747 / 3845 (45.44%) |

#### B. Not Supported Variants in MOAlmanac

| Category | Percent of all MOAlmanac Features |
|---|---|
| Sequence | 127 / 452 (28.10%) |
| Genotype/Haplotype | 0 / 452 (0.00%) |
| Fusion | 0 / 452 (0.00%) |
| Rearrangement | 35 / 452 (7.74%) |
| Epigenetic Modification | 0 / 452 (0.00%) |
| Copy Number | 23 / 452 (5.09%) |
| Expression | 11 / 452 (2.43%) |
| Gene Function | 8 / 452 (1.77%) |
| Region-Defined | 40 / 452 (8.85%) |
| Genome Feature | 0 / 452 (0.00%) |
| Transcript | 0 / 452 (0.00%) |
| Other | 12 / 452 (2.65%) |

**Supplemental Table 3: MOAlmanac Normalization Performance**. A. The number of variants within MOAlmanac that were successfully normalized or not supported. B. The number of not supported variants in MOAlmanac sorted by category.

#### Patient Sample Level Matching of GENIE Data

| | Protein | Genomic | Protein or Genomic |
|---|---|---|---|
| **All Knowledgebases** | 59.2 | 88.1 | 89.2 |
| **MOAlmanac** | 32.6 | 0 | 32.6 |
| **ClinVar** | 0 | 88.0 | 88.0 |
| **CIViC** | 58.3 | 0.7 | 58.6 |

**Supplemental Table 4: Percent of patient samples with at least one variant match for both protein and genomic variant representations.** MOA did not contain normalized genomic variants and ClinVar did not contain normalized protein variants, resulting in no matches for those respective categories.
